# Disparate and shared transcriptomic signatures associated with cortical atrophy in genetic bvFTD

**DOI:** 10.1101/2024.07.25.24310894

**Authors:** Ting Shen, Jacob W. Vogel, Vivianna M Van Deerlin, EunRan Suh, Laynie Dratch, Jeffrey S. Phillips, Lauren Massimo, Edward B. Lee, David J. Irwin, Corey T. McMillan

**Affiliations:** Penn Frontotemporal Degeneration Center, Department of Neurology, Perelman School of Medicine, University of Pennsylvania, Philadelphia, PA, USA; Department of Clinical Sciences Malmö, SciLifeLab, Lund University, Lund, Sweden; Center for Neurodegenerative Disease Research, Department of Pathology and Laboratory Medicine, Perelman School of Medicine, University of Pennsylvania, Philadelphia, PA, USA

**Keywords:** behavioral variant frontotemporal dementia, transcriptomics, cortical thickness, partial least squares regression, synaptic density, pathology

## Abstract

Cortical atrophy in behavioral variant frontotemporal degeneration (bvFTD) exhibits spatial heterogeneity across genetic subgroups, potentially driven by distinct biological mechanisms. Using an integrative imaging-transcriptomics approach, we identified disparate and shared transcriptomic signatures associated with cortical thickness in *C9orf72*, *GRN* or *MAPT*-related bvFTD. Genes associated with cortical thinning in *GRN*-bvFTD were implicated in neurotransmission, further supported by mapping synaptic density maps to cortical thickness maps. Previously identified genes linked to TDP-43 positive neurons were significantly overlapped with genes associated with *C9orf72*-bvFTD and *GRN*-bvFTD, but not *MAPT*-bvFTD providing specificity for our associations. *C9orf72*-bvFTD and *GRN*-bvFTD shared genes displaying consistent directionality of correlations with cortical thickness, while *MAPT*-bvFTD displayed more pronounced differences in transcriptomic signatures with opposing directionality. Overall, we identified disparate and shared genes tied to regional vulnerability with increased biological interpretation including overlap with synaptic density maps and pathologically-specific gene expression, illuminating intricate molecular underpinnings contributing to heterogeneities in bvFTD.

## Introduction

Behavioral variant frontotemporal dementia (bvFTD) is characterized by a progressive deterioration of personality and social behavior, and often accompanied by cognitive impairments^1^. While the majority of individuals with bvFTD have apparently sporadically illness, a subset of bvFTD have pathogenic variants in genes associated with autosomal dominant inheritance of FTD, with a greater likelihood of a genetic etiology in the presence of family history of FTD or related conditions. Approximately 15-20% individuals with bvFTD have an identifiable genetic etiology^2^, with the most prevalent genetic causes including repeat expansions of *C9orf72*, pathogenic variants in *GRN* and *MAPT* genes^3^. Genetic forms are often linked to distinct clinical presentations, underlying pathologies, and neuroanatomical patterns in comparison to apparently sporadic bvFTD, which complicate the disease landscape.

Cortical degeneration is a common neuroanatomical manifestation in bvFTD, primarily affecting the frontal and anterior temporal lobe. Distinct brain atrophy patterns have been observed among different genetic forms. BvFTD with repeat expansions in *C9orf72* (*C9orf72*-bvFTD) exhibits a higher degree of atrophy in parietal and occipital lobes, as well as the lateral inferior frontal lobe and cerebellum, compared with bvFTD carrying *MAPT* pathogenic variants (*MAPT*-bvFTD) and apparently sporadic bvFTD^4,5^. Compared to *C9orf72*-bvFTD, *MAPT*-bvFTD demonstrates more pronounced atrophy in the anterior temporal lobes, while bvFTD carrying *GRN* pathogenic variants (*GRN*-bvFTD) shows greater grey matter loss in inferior temporal and parietal lobes^4,5^. Moreover, frontotemporal lobar degeneration (FTLD) with *GRN* pathogenic variants experience faster progression of atrophy than *C9orf72*-FTLD and *MAPT*-FTLD^6^. Distinct atrophy patterns across various forms of genetic bvFTD may be shaped by their different biological mechanisms.

Different genetic forms of bvFTD may exhibit distinct and shared molecular signatures that influence the disease presentation and progression. Genetic variants may lead to dysregulated molecular pathways related to the mutated genes. For instance, the dysfunction of C9orf72 protein may interrupt the regulation of endosomal trafficking, autophagy, lysosomal and microglial functions, and lead to neurodegeneration^7^. The *MAPT* pathogenic variants give rise to a cascade of events that cause the dysregulation of synaptic, neuronal, and lysosomal functions^8^. Within the frontal cortex of *GRN*-FTD, there are upregulated genes associated with TGF-beta signaling and cell communication, while downregulated genes are involved in calcium signaling^9^. Additionally, another study further analyzed transcriptomics data in cerebellum, frontal cortex, and hippocampus, which revealed that dysregulated genes in *GRN*-FTD primarily centered on Wnt signaling pathway^10^. We hypothesize that the regional distribution of gene expression may contribute to selective vulnerability of regional neurodegeneration, giving rise to a variety of distinct neuroanatomic patterns observed in genetic bvFTD. Therefore, it is crucial to consider each genetic form of bvFTD individually to better understand the complex genetic underpinnings of bvFTD.

Here, we sought to bridge these gaps by relating transcriptomic data from the Allen Human Brain Atlas (AHBA) to macroscale structural neuroimaging in various forms of genetic bvFTD (*C9orf72*, *GRN*, and *MAPT*), relative to individuals with apparently sporadic bvFTD. Our goal was to explore potential molecular and cellular pathways that might explain regional vulnerability to neurodegeneration due to genetic pathogenic variants. The study framework is summarized in Figure 1.

**Figure 1.**
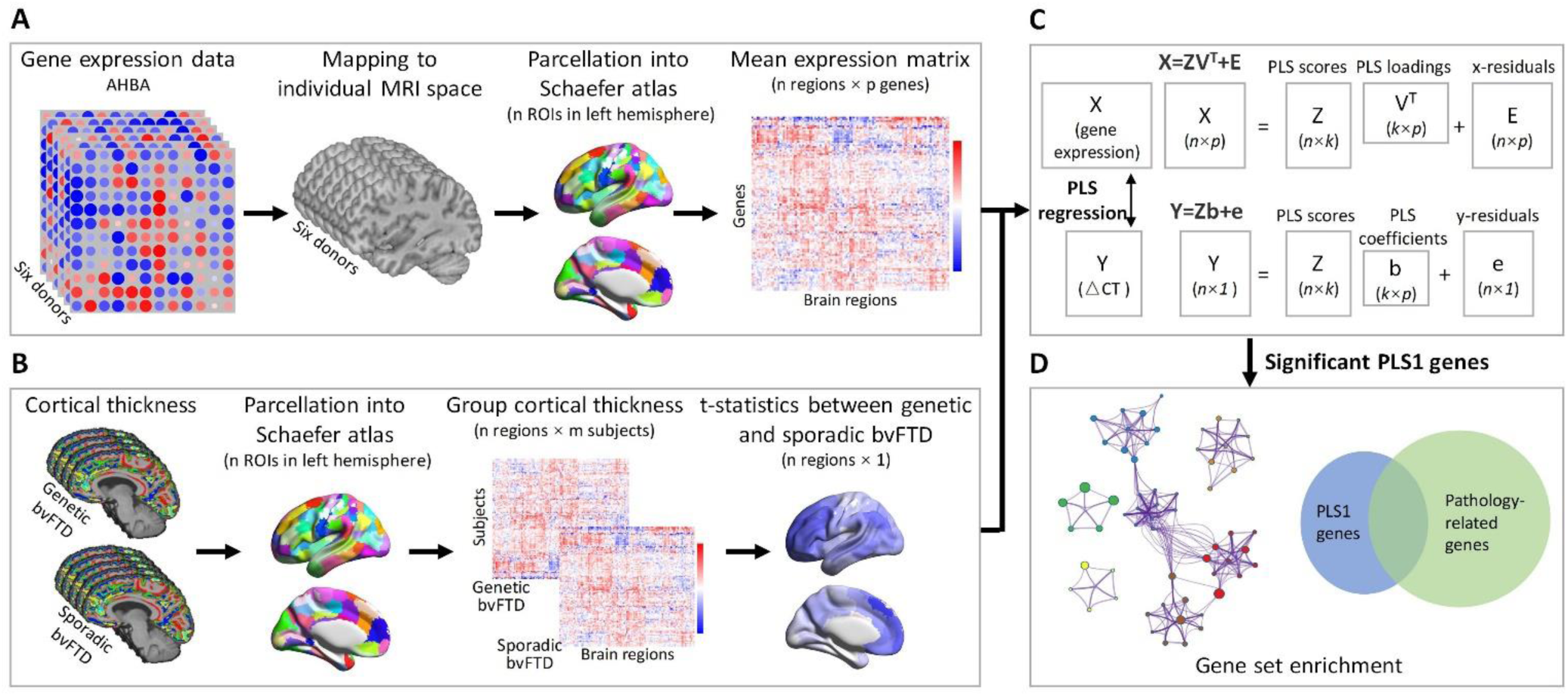
Overview of study pipeline. (A) Regional gene expression data extracted from the AHBA in 100 regions (left hemisphere only) across the 6 donors. (B) Cortical thickness data extracted from regions in the same atlas. The cortical thickness signatures were computed as t-statistic values compared between individuals with genetic and apparently sporadic bvFTD. (C) PLS regression was used to identify imaging transcriptomic associations. Genes with significant association were retained for subsequent analyses. (D) Functional enrichment analyses were conducted on genes with significantly loading on the PLS1, including functional enrichment and pathology enrichment analyses.

## Results

### Demographics and clinical characteristics

Table 1 displays the demographic and clinical features of 173 individuals with bvFTD, comprised of 117 individuals with apparently sporadic disease, 32 with *C9orf72* repeat expansions, 11 with pathogenic variants in *GRN*, and 13 with pathogenic variants in *MAPT*. Individuals with *MAPT*-bvFTD tended to be younger than other genetic forms or apparently sporadic cases. The individuals with *GRN*-bvFTD had a shorter disease duration compared to those with apparently sporadic bvFTD.

**Table 1.** Demographic and clinical characteristics of the study population.

|  | Apparently<br>sporadic (n = 117) | C9orf72 (n = 32) | GRN (n = 11) | MAPT (n = 13) | Missing<br>data |
| --- | --- | --- | --- | --- | --- |
| Age at MRI (years) | 62.3 (8.6) | 62.7 (7.1) | 62.1 (4.4) | 54.0 (7.6)<sup>a</sup> | 0.0% |
| Sex (male%) | 72 (61.5%) | 20 (62.5%) | 6 (54.6%) | 6 (46.2%) | 0.0% |
| Disease duration (years)<sup>b</sup> | 3.9 (2.7) | 3.8 (2.7) | 2.2 (1.2)<sup>a</sup> | 3.6 (2.6) | 0.0% |
| MMSE | 24.1 (5.7) | 23.9 (5.6) | 21.3 (7.4) | 22.8 (5.2) | 12.1% |
| PBAC total | 57.9 (15.6) | 64.5 (13.2) | 55.0 (16.0) | 65.3 (13.7) | 31.2% |

Data are presented as mean (standard deviation) for the continuous variables, and as number (frequency) for the categorical variables.
<sup>a</sup> indicates significant differences compared with the individuals with apparently sporadic bvFTD.
<sup>b</sup> Disease duration is the interval between symptom onset and the time of scan.
bvFTD, behavioral variant frontotemporal dementia; MMSE, Mini-Mental Status Examination; PBAC, Philadelphia Brief Assessment of Cognition

### Cortical thickness signatures of different genetic forms of bvFTD

To identify cortical thickness signatures specific to genetic bvFTD, we compared the cortical thickness between each genetic group and apparently sporadic bvFTD (Figure S1). In comparison to the apparently sporadic bvFTD, individuals with *C9orf72*-bvFTD showed relatively spared cortical thickness in anterior and inferolateral temporal. *GRN*-bvFTD exhibited more extensive global reductions in cortical thickness relative to apparently sporadic bvFTD, most predominant in inferior frontal cortex and parietal regions including precuneus and posterior cingulate cortex. Relative to apparently sporadic bvFTD, *MAPT*-bvFTD was associated with globally reduced cortical thickness with more predominantly reduced cortical thickness in insula, motor cortex, and dorsolateral prefrontal cortex.

### Gene expression patterns associated with cortical thickness signatures

We used a PLS regression model to examine the relationship between gene expression and anatomical distribution of cortical thickness signatures across brain regions. The PLS1 was the optimal representation of the covariance of high-dimensional data, capturing the maximum gene expression variance across cortical regions in AHBA. Positive weights (PLS1+) represented that these genes were overexpressed in regions where cortical thickness increased, while negative weights (PLS1-) mean that expression levels of those genes increased in regions where the cortical thickness decreased. This procedure was performed separately for each genetic form of bvFTD. All significant genes were shown in Table S1.

For *C9orf72*-bvFTD, the PLS1 component explains 26.6% of the variance in cortical thickness signature, which is greater than chance (*p_boot_* = 0.001; Figure 2A). The weighted gene expression map of the PLS1 component exhibited a gradient of gene expression levels, ranging from higher gene expression in left-predominant anterior temporal, insular, and anterior cingulate regions and lower gene expression associated with the more spared parietal-occipital cortex (Figure 2B). The gene expression level, reflected by the PLS1 score in each region correlated with the regional t-statistic map of cortical thickness for the *C9orf72*-bvFTD relative to the apparently sporadic bvFTD (Figure 2C). To identify significant contributors, all genes were ranked by PLS1 weights, and those with variable importance in projection (VIP) > 1^11^ and *p-values* < 0.05 were considered as significant genes (Figure 2D). We identified 191 PLS1+ and 175 PLS1- genes for *C9orf72*-bvFTD (Figure 2E). In *GRN*-bvFTD, we observed that the PLS1 component explained 25.1% of the variance in cortical atrophy pattern (*p_boot_* = 0.08; Figure 2A). Similarly, the PLS1 weighted gene expression map also displayed a pattern of higher gene expression in anterior regions and lower gene expression in posterior regions (Figure 2B), which was spatially correlated with the regional cortical thickness t-statistic map (Figure 2C). We identified 134 PLS1+ and 363 PLS1- genes for this group (Figure 2E). Regarding the *MAPT*-bvFTD, cortical atrophy was accounted by PLS1 to an extent of approximately 21.0% (*p_boot_* = 0.36; Figure 2A). Notably diverging from the patterns observed in *C9orf72*-bvFTD and *GRN*-bvFTD, a distinct feature of this group was the presence of a posterior-anterior gradient in the distribution of gene expression (Figure 2B). We identified 321 PLS1+ and 233 PLS1- genes for *MAPT*-bvFTD (Figure 2E).

**Figure 2.**
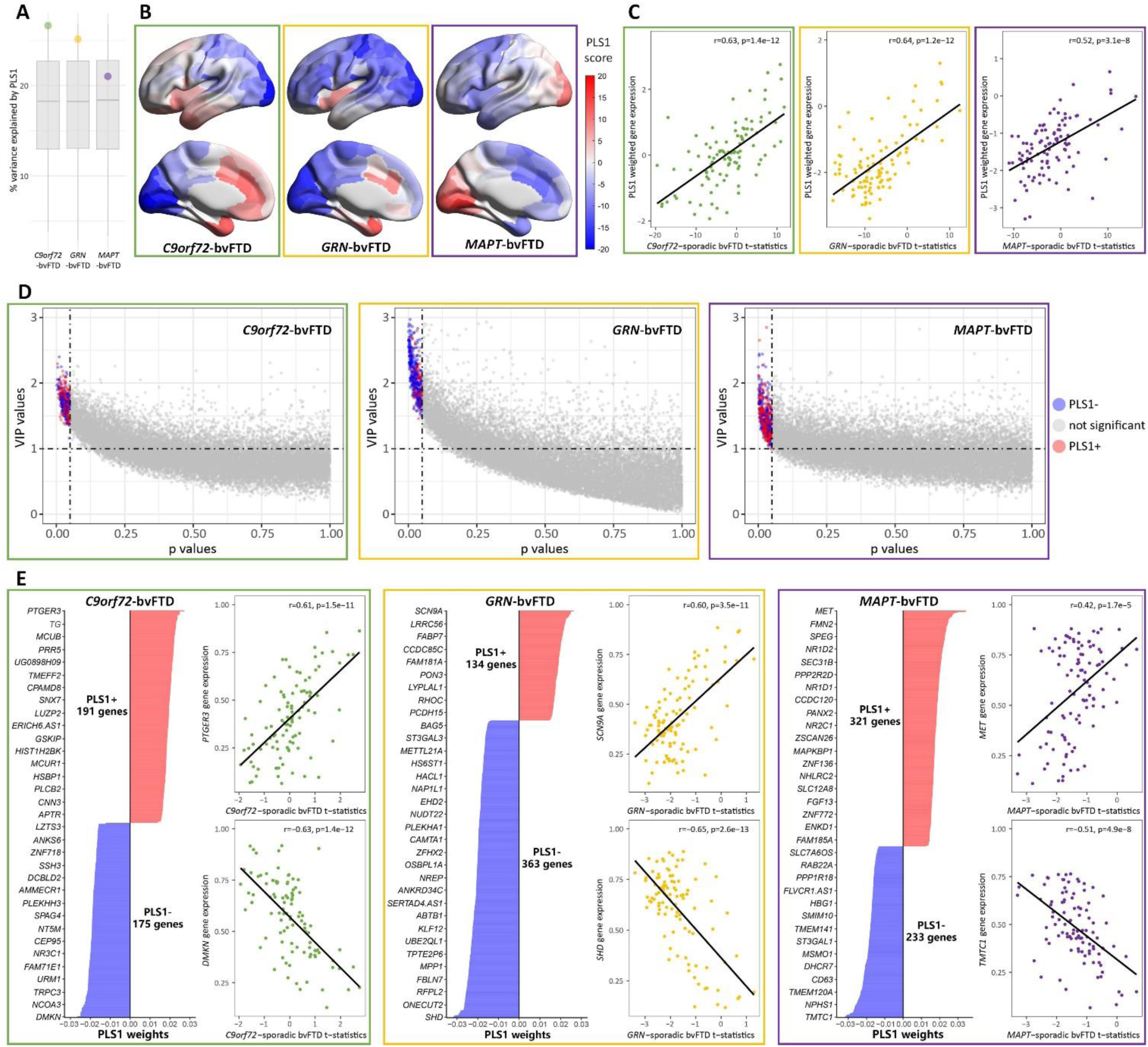
Gene expression patterns associated with cortical thickness signatures. (A) The percentage of the variance in cortical thickness signature explained by PLS1. The boxplots show the null distribution of variance explained by PLS1, each dot represent the observed value. (B) PLS1 weighted gene expression of each genetic form of bvFTD. Darker red indicates higher PLS1 scores, whereas darker blue corresponds to lower scores. (C) Significantly positive correlations between PLS1 weighted gene expression and anatomical map of regional cortical thickness signatures. (D) Genes with a VIP score > 1, and bootstrapping *p value* < 0.05 were selected as top-ranked genes. Blue dots represent genes with expression levels negatively correlated with cortical thickness, red dots represent genes with expression levels positively correlated with cortical thickness, and gray dots represent genes with no significant correlation. (E) Bar plots display genes ranked by their PLS1 weights. The weights are coefficients of the linear combination of the original variables used to predict the response variables in the PLS regression model. Scatter plots show correlations between the anatomical map of regional cortical thickness signatures and gene expression level of the top positively or negatively weighted genes.

### Functional enrichment analyses

We performed functional enrichment analyses on gene sets categorized as PLS1+ and PLS1- separately (Table S2). The PLS1-/+ gene sets associated with *C9orf72*-bvFTD did not exhibit statistically significant enrichments. However, the PLS1+ gene list showed connections with biological processes including “neurotransmission and synaptic signaling”, “neural development and structure”, and “perception”. The PLS1- genes of *GRN*-bvFTD were significantly enriched for neural-related terms, particularly those involved in “neuronal membrane components”, “regulation of synaptic membrane potential” (Figure 3A). In addition, this list was also associated with biological processes related to “ion channel-related transport” (Figure 3A). The PLS1+ genes of *GRN*-bvFTD were significantly enriched in “circadian entrainment” (Figure 3B). These PLS-/+ gene lists of *GRN*-bvFTD both have connections with “neurotransmission and synaptic signaling”, despite not being statistically enriched. A notable observation is that both PLS1- and PLS1+ gene lists of *GRN*-bvFTD were linked to gamma-aminobutyric acid-ergic (GABAergic) synapse and neurotransmission. In addition, the PLS1- genes were associated with dopaminergic synapse, while the PLS1+ genes were linked to glutamatergic, cholinergic, and serotonergic synapses. The PLS1- genes of *MAPT*-bvFTD were enriched in biological terms related to “cholesterol biosynthesis and metabolism”, and “enzyme activities and metabolism”, which were involved in various systems, not specific to nervous system (Figure 3C). Additionally, this gene set tended to associated with pathways related to Parkinson’s disease (PD). While not reaching statistical significance, the PLS1+ genes of *MAPT*-bvFTD was also associated with “circadian rhythm” and “regulation of behavior”.

**Figure 3.**
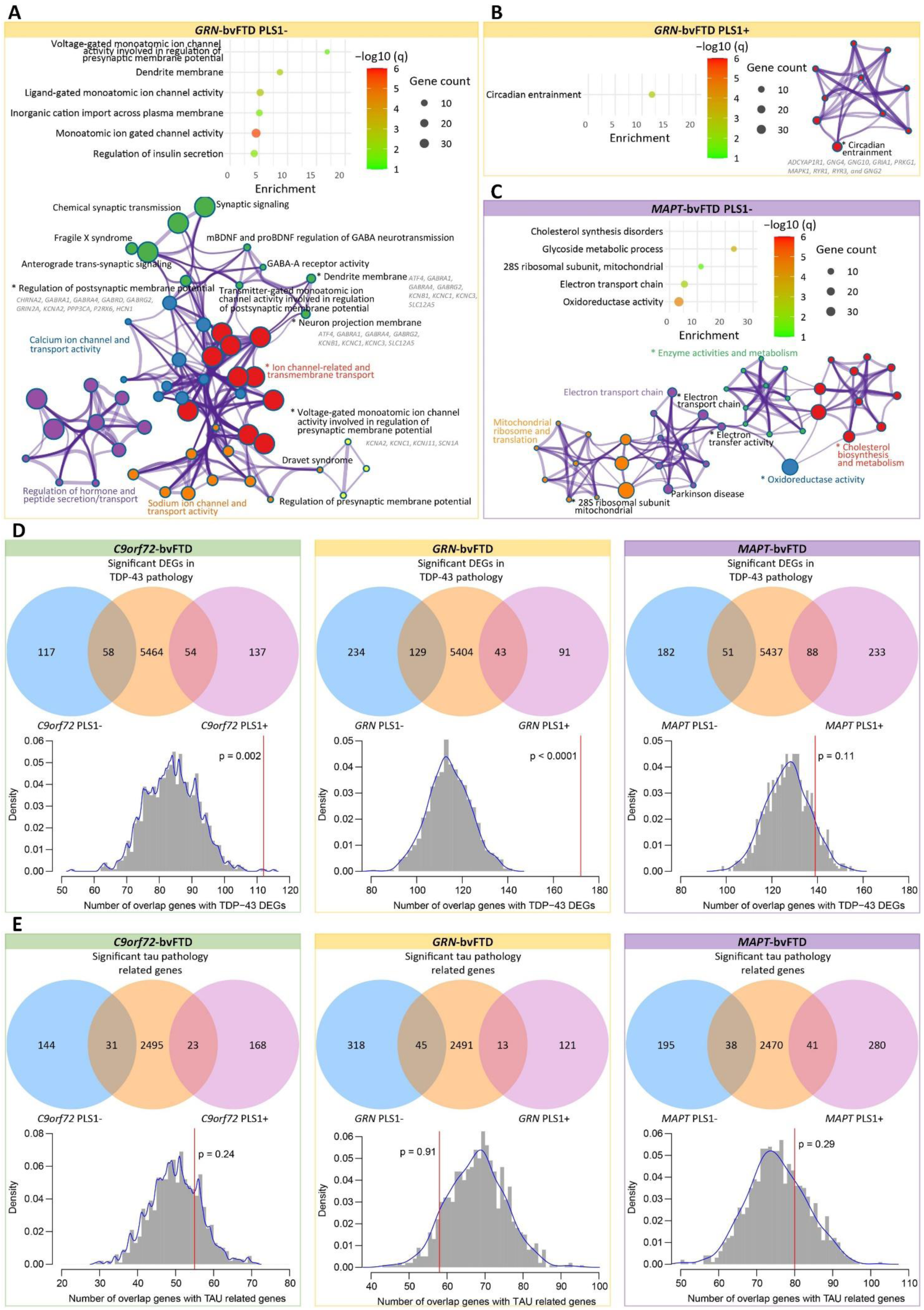
Functional enrichment and Pathology-related enrichment analyses. (A and B) Results of functional enrichment of PLS1- (A) and PLS1+ (B) genes associated with cortical thickness signatures of *GRN*-bvFTD. (C) Results of functional enrichment of PLS1- genes associated with cortical thickness signatures in *MAPT*-bvFTD. All significant resultant terms were clustered into groups based on their similarities. The bubble plot shows the subset of representative ontology terms for each enriched cluster. The bubble size represents the overlapping gene count between the PLS1 gene set and genes involved in each ontology term. The color bar represents the logarithmically transformed BH-FDR-corrected *q values*. Metascape enrichment network visualization showing the intra-cluster and inter-cluster similarities of enriched terms. Nodes are colored to reflect their cluster memberships. * indicates that the biological processes were significant. Genes that involved in these significant terms were listed in the figure. (D-E) The overlap between PLS1 gene lists associated with each genetic form of bvFTD and TDP-43 pathology-related DEGs (D) and tau pathology-related genes (E). Venn diagrams (upper) showing the overlap between groups. Histograms (bottom) display the null distribution, depicting the number of overlapped genes between PLS1 genes and pathologies related genes. The red line represents the observed number of overlapped genes.

### Pathology-related gene enrichment analyses

To investigate that whether these PLS1-/+ genes are implicated in neuropathology of FTD, we calculated the number of overlapping genes between our gene lists and previously implicated TDP-43 and tau pathology-related gene lists. In a gene list comprised of DEGs observed in fluorescent-activated cell sorting (FACS) sorted TDP- 43 positive compared to TDP-43 negative neurons^12^, our findings revealed that PLS1 genes associated with *C9orf72*-bvFTD and *GRN*-bvFTD were enriched in these genes previously linked to TDP-43 pathology, whereas gene sets linked to *MAPT*-bvFTD did not show significant enrichment (Figure 3D). Upon investigating the intersection of genes associated with TDP-43 pathology, we found that among the PLS1 genes associated with *C9orf72*-bvFTD, there were 112 genes, while there were 172 genes associated with *GRN*-bvFTD, and 139 genes linked to *MAPT*-bvFTD (Table S3). Permutation testing relative to 1,000 random subsets of genes confirmed this overlap significantly differed from chance, suggesting our indirect associations of gene expression with neuroimaging features converge with known TDP-43 pathogenesis. Notably, these shared genes in *C9orf72*-bvFTD and *GRN*-bvFTD groups significantly encompass crucial components involved in neural pathways of “neurotransmission and synaptic signaling” (Table S4).

In the analysis of tau pathology in PSP, a previous study identified a gene set significantly associated with the overall burden of tau pathology^13^. However, we did not observe significant enrichment of the three genetic forms in this tau pathology-related gene set (Figure 3E). When examining the overlap with the tau pathology-related gene set, there were 55 genes in the PLS1 gene list associated with *C9orf72*- bvFTD, 58 genes in the PLS1 gene list associated with *GRN*-bvFTD, and 80 genes in the PLS1 gene list associated with *MAPT*-bvFTD (Table S3). While not reaching statistical significance, these shared genes were mainly associated with biological processes that were involved in various systems, such as “transcriptional regulation and protein processes”, “cellular processes and organization”, and “ion transport and signaling” (Table S4), rather than pathways highly specific to nervous system.

### Mapping to synaptic density maps

After identifying gene sets associated with “neurotransmission and synaptic signaling”, we extracted density maps of five neurotransmitter systems from published studies, and subsequently correlated them with cortical thickness t-statistic map of each bvFTD genetic form (Figure 4A). *C9orf72*-bvFTD t-statistic map exhibited positive correlations with dopaminergic (D2 receptor) and serotonergic (5-HT1_A_ receptor) neurotransmitter densities, while showing negative correlations with GABAergic neurotransmitter receptor density. *GRN*-bvFTD t-statistic map showed positive correlations with dopaminergic neurotransmitters, while showing either positive or negative correlations with different receptors of other neurotransmitter systems. *MAPT*-bvFTD t-statistic map was positively correlated with GABAergic neurotransmitter receptor density and negatively related to cholinergic neurotransmitter density. For dopaminergic neurotransmitters, it is positively correlated with D1 receptor density and negatively correlated with D2 receptor density. Furthermore, dominance analysis, which consider the correlations between variables, were conducted to evaluate the importance of the 15 neurotransmitter receptors/transporters in relation to cortical atrophy in different genetic form of bvFTD (Figure 4B). The serotonergic (5-HT1_A_ receptor) and GABAergic neurotransmitter systems were the most important variables relating to cortical thickness in *C9orf72*-bvFTD. In *GRN*-bvFTD, the serotonergic (5-HT1_A_ and 5-HTT receptors) and dopaminergic (dopamine transporter) neurotransmitter systems were the most important variables. In *MAPT*-bvFTD, dopaminergic (D_1_ and D_2_ receptors) and cholinergic (α_4_β_2_ receptor) neurotransmitter systems showed the highest importance in relation to cortical thickness.

**Figure 4.**
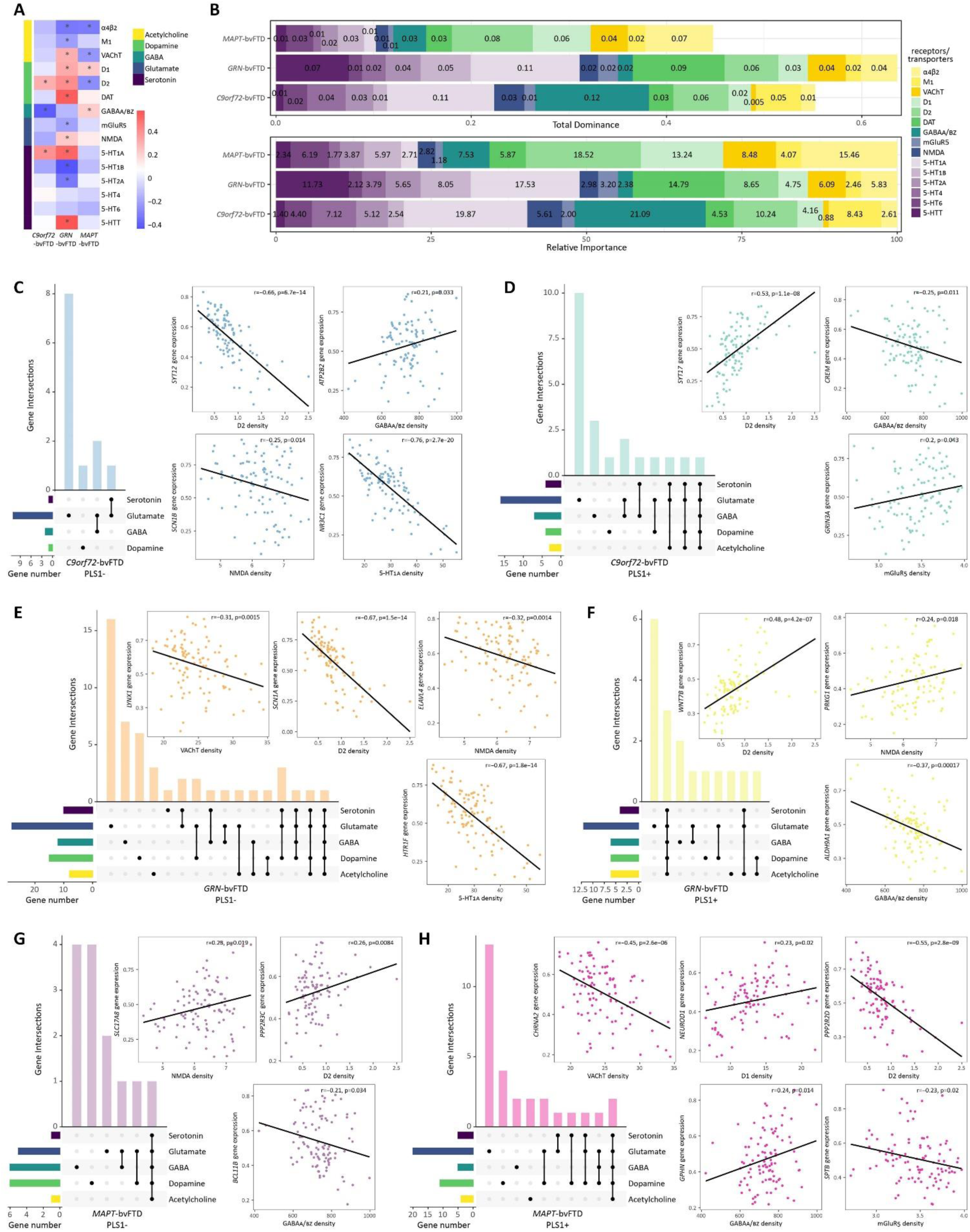
Mapping to synaptic density maps. Correlations between cortical thickness t-statistic maps and neurotransmitter receptor/transporter density maps. * indicates that the correlation is significant. (B) Results of dominance analysis. Upper bar plot displays the importance of 15 neurotransmitter receptor/transporter densities in predicting cortical atrophy. Lower bar plot illustrates the relative importance of each variable. (C-H) Upset plots illustrate genes involved in five neurotransmitter systems. Horizontal bars show the total number of genes involved in each neurotransmitter system. Bottom matrix indicates neurotransmitter systems involved in each intersection. Filled dot indicates the presence of a system in that intersection, and connected dots indicate a combination of systems in that intersection. Scatter plot shows the correlation between the expression map of a certain gene and a neurotransmitter density map.

Membership analyses revealed that every PLS1 gene set contains genes belonging to terms of these neurotransmitters. Their gene expression maps were then correlated with the neurotransmitter receptor/transporter density maps (Figure 4C-H and Figure S2). Higher proportion of genes in *MAPT*- bvFTD PLS1- list were involved in dopaminergic and GABAergic terms, while other gene lists were more related to glutamatergic system. For *GRN*-bvFTD, the terms identified in membership analysis involved five neurotransmitters, all significantly enriched in PLS1-/+ gene lists (Figure S3). These findings are consistent with the significant correlations observed between its cortical thickness t-statistic map and neurotransmitter maps, further validating that cortical atrophy associated with pathogenic variants in *GRN* is more closely related to mechanisms involving neurotransmission and synaptic signaling.

### Shared genes across genetic forms of bvFTD

Despite different genetic sources, bvFTD presents as a relatively homogenous syndrome, suggesting that there may be shared genetic factors influencing brain structures across various genetic forms. Therefore, we further identified shared transcriptomic signatures associated with cortical atrophy across three genetic forms. Figure 5A-C showed the number of overlapping genes within gene sets positively and negatively associated with bvFTD-related atrophy. *C9orf72*-bvFTD and *GRN*-bvFTD had several overlapping genes with shared directionality. Specifically, genes with positive weights in *C9orf72*-bvFTD also displayed positive weights in *GRN*-bvFTD. Thus, in these two genetic forms, these shared genes were commonly expressed in regions more spared by cortical atrophy. Similarly, the shared genes with negative weights in *C9orf72*-bvFTD also displayed negative weights in *GRN*-bvFTD, suggesting that these genes were commonly expressed in regions more vulnerable to brain atrophy in both genetic forms. *MAPT*-bvFTD showed several overlapping genes with the other two forms, all with opposing directionality. This indicates that these shared genes had opposite correlations with cortical thickness in *C9orf72*-bvFTD and *GRN*-bvFTD compared to *MAPT*-bvFTD.

**Figure 5.**
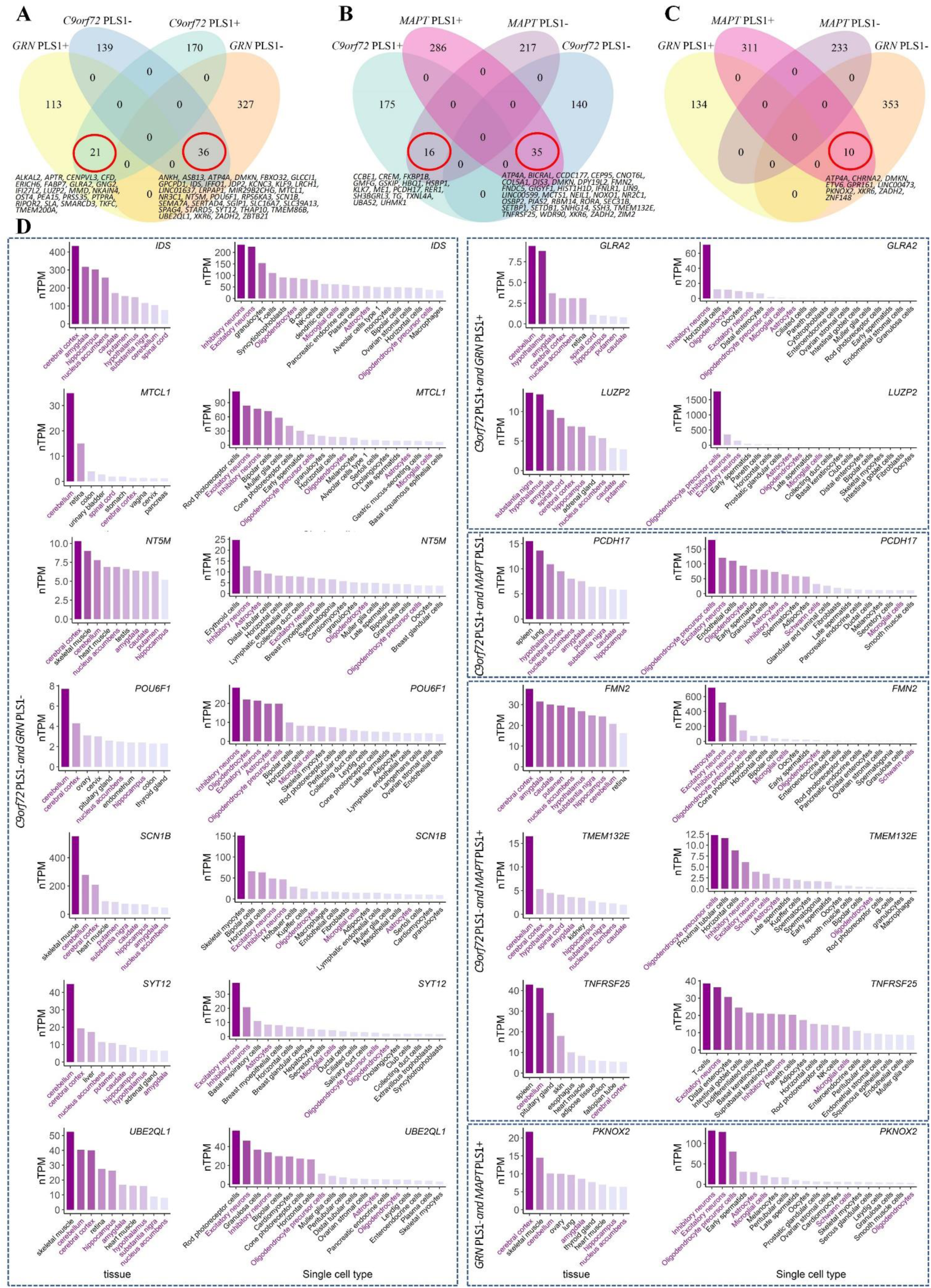
**Shared genes across genetic forms of bvFTD.** (A) Venn diagrams showing the genes that associated with cortical thickness were shared between *C9orf72*-bvFTD and *GRN*-bvFTD. (B) Shared genes between *C9orf72*-bvFTD and *MAPT*-bvFTD. (C) Shared genes between *GRN*-bvFTD and *MAPT*-bvFTD. (D) STRING interaction diagram of the overlapped genes. Each node denotes a gene, edges between nodes indicate interactions between protein products of the corresponding genes, with the line thickness indicating the strength of interactions. Genes are colored according to the functional pathways that these genes involved. The genes circled in red represent the overlapped genes that we identified, while the genes without circles are predicted functional partners provided by STRING. (E) The genes expressed in multiple tissues and cell types. Only genes specifically enriched in both brain tissues and neural cells were shown.

Afterwards, we constructed PPI networks using STRING database (Figure 5D). Among the overlapping genes between *C9orf72*-bvFTD and *GRN*-bvFTD, G protein subunit gamma 2 (*GNG2*) was involved in synaptic signaling and circadian entrainment. The potassium voltage-gated channel subfamily C member 3 (*KCNC3*) and ribosomal protein S6 kinase alpha-3 (*RPS6KA3*) genes may contribute to other aspects of neuronal system. The SWI/SNF-related, matrix-associated, actin dependent regulator of chromatin, subfamily d, member 3 (*SMARCD3*) gene encodes a protein belonging to SWI/SNF family, involving in regulating gene transcription through chromatin remodeling. The shared genes with opposing directionality in *GRN*-bvFTD and *MAPT*-bvFTD included the cholinergic receptor nicotinic alpha 2 subunit (*CHRNA2*), G protein-coupled receptor 161 (*GPR161*), and zinc finger protein 148 (*ZNF148*) that were associated with abnormality of nervous system. Additionally, the ETS Variant Transcription Factor 6 (*ETV6*) gene was linked to genes that involved in synaptic signaling and axon guidance. When examining the overlapping genes with opposing directionality in *C9orf72*-bvFTD and *MAPT*-bvFTD, the shared biological pathways primarily involved genes that are directly or indirectly associated with RNA degradation, ribosome function, and cellular responses to stress.

Among the genes that overlapped, the glycine receptor alpha 2 (*GLRA2*), leucine zipper protein 2 (*LUZP2*), iduronate 2-sulfatase (*IDS*), microtubule crosslinking factor 1 (*MTCL1*), 5’,3’-nucleotidase, mitochondrial (*NT5M*), POU class 6 homeobox 1 (*POU6F1*), sodium voltage-gated channel beta subunit 1 (*SCN1B*), synaptotagmin 12 (*SYT12*), ubiquitin conjugating enzyme E2 Q family like 1 (*UBE2QL1*), protocadherin 17 (*PCDH17*), formin 2 (*FMN2*), transmembrane protein 132E (*TMEM132E*), TNF receptor superfamily member 25 (*TNFRSF25*), and PBX/knotted 1 homeobox 2 (*PKNOX2*) genes, were specifically enriched in brain and neural cells, as indicated by GTEx database and single cell type expression maps in HPA database. Additionally, there were 17 other genes that showed specific enrichment in brain neural cells, while 14 genes displayed specific enrichment in brain tissues (Figure S4).

## Discussion

This study employed an integrative imaging-transcriptomic approach to uncover the transcriptomic signatures associated with cortical thinning across various genetic subgroups of bvFTD. We observed distinct patterns of cortical thinning in each genetic form of bvFTD compared to apparently sporadic bvFTD. Subsequently, we utilized a PLS regression model to identify genes whose expression levels across brain regions exhibited strong correlations with cortical thinning. Our findings suggested the presence of distinct molecular mechanisms specific to different genetic forms, and shared molecular mechanisms contributing to cortical remodeling in different genetic forms.

Diverse but overlapping brain atrophy patterns have been identified among various genetic forms of bvFTD^4^. In our study, individuals with *C9orf72*-bvFTD exhibited higher cortical thickness in temporal lobe compared to those with apparently sporadic bvFTD. This observation aligns with previous findings indicating that individuals with *C9orf72* repeat expansions rarely present with semantic variant primary progressive aphasia (svPPA), which is a temporal-predominant syndrome^3,14^. Moreover, FTLD-TDP type A and type B are predominant subtypes that associated with *C9orf72*, were more spared in temporal cortex^15,16^. In line with previous findings of temporal and orbitofrontal lobe atrophy in *MAPT*-FTD^17^, we also observed greater grey matter loss in temporal pole and prefrontal cortex, alongside other regions such as somatomotor cortex, frontal operculum insula, and parietal lobe in *MAPT*-bvFTD. On the other hand, those with *GRN*-bvFTD exhibited more extensive cortical atrophy, affecting various brain regions including prefrontal, temporal, somatomotor, visual, posterior cortices, precuneus and posterior cingulate. Individuals with *GRN*-bvFTD demonstrated later disease onset, shorter disease duration, and faster atrophy progression, suggesting that they may have a prolonged latent phase, however once it reaches a certain stage, the disease progresses rapidly. Previous study also reported that *GRN*-FTLD experienced faster atrophy progression compared to other genetic forms^6^. These observed patterns may not necessarily reflect the overall clinical phenotype of bvFTD, but could be solely attributed to the pathogenic variants rather than being driven by the clinical features of bvFTD.

Using a PLS regression model, we identified significant transcriptional associations with neuroimaging. The PLS1 component, capturing the maximum proportion of covariance between gene expression and cortical thickness signatures, provided valuable insights into the gene expression patterns colocalizing with cortical thickness signatures in each genetic subgroup. Our analysis yielded lists of PLS1 genes that negatively or positively linked to cortical atrophy for each genetic form. These spatial gene expression patterns could potentially constitute molecular foundation of disease-related cortical remodeling. Distinct gene transcripts specific to each genetic form shed light on the molecular mechanisms driving cortical remodeling in different genetic forms of bvFTD. This to some extent reveals the molecular mechanisms that underlie the heterogeneity of observed neuroanatomical changes across the genetic bvFTD population.

Extensive frontotemporal synaptic loss has been documented in bvFTD, correlating with disease severity^18^. BvFTD is a major clinical subtype of FTLD^1^, associated with alterations in various neurotransmitter systems, including glutamatergic, GABAergic, dopaminergic, and serotonergic systems^19^, constituting a fundamental molecular mechanism underlying bvFTD. Among genes that positively associated with cortical thickness signature of *C9orf72*-bvFTD, there were certain genes holding notable functional importance in “neurotransmission and synaptic signaling”, with a specific emphasis on GABAergic systems. Additionally, there also existed genes related to “neural development and structure”, such as the cytoplasm of neuron projections (axons). As widely recognized, repeat expansions in *C9orf72* triggers abnormal RNA-processing events, crucially impacting the organization of synapses and their cytoskeletal components, as well as modulating synaptic strength and function^20^. Similarly, within the genes associated with cortical atrophy in *GRN*-bvFTD, there were genes connecting to “neurotransmission and synaptic signaling”. Both the negatively and positively associated gene sets encompassed genes related to GABAergic system. Furthermore, negatively associated genes were related to dopaminergic synapse, whereas the positively associated genes were linked to glutamatergic, cholinergic, and serotonergic synapses. Our findings align with previous studies, as pathogenic variants in *GRN* may impact synaptic function, such as impairments in synaptic connectivity, plasticity, and transmission^21^. In general, these genes associated with *C9orf72*-bvFTD and *GRN*-bvFTD may both influence neurotransmission and synaptic signaling, contributing to cortical remodeling. However, the distinction might lie in the specific aspects of the synaptic systems they affect.

Most of the genes we identified that associated with *MAPT*-bvFTD were not specifically link to nervous system. Just as the haplotype of *MAPT* gene is linked to PD^22^, several genes we have identified could also potentially be involved in biological processes related to PD. In addition to impairments in behavior, cognition, and movement, notable changes have also occurred in sleep patterns or circadian function in bvFTD^23^. Circadian dysregulation appears to occur before the onset of cognitive symptoms and tends to deteriorate with disease progression of FTD^24^. Neurodegenerative diseases are linked to pathological disruptions in circadian and sleep regulatory networks that encompass brain regions such as hypothalamus, basal forebrain, and mesial temporal lobe^24^. In our study, certain genes positively associated with *MAPT*-bvFTD were involved in “circadian”, so as certain genes positively associated with *GRN*-bvFTD. This observation aligns with a previous study which reported a greater tau burden compared to TDP-43 burden in locus coeruleus, a region implicated in circadian functions^25^. From a spatial transcriptomics perspective, *C9orf72* may also be connected to circadian-related pathways. This is evidenced by our discovery that several genes associated with *C9orf72*-bvFTD were specifically expressed in cholinergic neurons in basal forebrain and hypocretinergic neurons in hypothalamus. Notably, these regions and pathways play pivotal roles in regulation of circadian system^26^. Furthermore, striking sleep disruption has been particularly reported in individuals with *C9orf72*-FTD^24^. Proteinopathies such as tauopathy have the potential to disrupt circadian rhythm. Conversely, circadian dysregulation also influences the occurrence and progression of these pathologies^27,28^. Circadian rhythmicity also involves the regulation of glymphatic system, a brain waste clearance system responsible for eliminating proteins and other solutes from interstitial fluid^29^. Circadian dysregulation may potentially lead to glymphatic system failure, impeding clearance of abnormal protein accumulations such as TDP-43 and tau proteins. Glymphatic dysfunction could serve as a common pathway towards various forms of dementia^29^. Thus, this shared involvement in circadian-related pathways may represent common pathological regulators of these genetic forms. However, specific genes within these pathways vary due to distinct genetic backgrounds.

Previous studies had shown widespread involvement of cholinergic, dopaminergic, serotonergic, and glutamatergic neurotransmissions across various genetic subgroups of FTD^30^. By mapping different neurotransmitter systems to cortical structure, we further confirmed that genes associated with brain atrophy in *GRN*-bvFTD were significantly related to synaptic signaling. This involvement spans multiple neurotransmitter systems, as reported in functional enrichment analyses, thereby reinforcing the role of various neurotransmitter pathways in cortical atrophy in *GRN*-bvFTD. For *C9orf72*-bvFTD, GABAergic signaling pathways contributed the most in predicting brain atrophy, supported by findings that genes associated with GABAergic signaling are impacted in *C9orf72*-FTLD^31^. In *MAPT*-bvFTD, cholinergic, dopaminergic and GABAergic neurotransmitters are important contributors in predicting brain atrophy. Individuals with pathogenic variants in *MAPT* gene were observed with dopaminergic dysfunction^32^. *MAPT* pathogenic variants may trigger alterations in GABAergic signaling and synaptic function, leading to pathogenesis of tauopathies^33^. These observations further highlight the critical role of neurotransmission and synaptic signaling in shaping brain structure, particularly in *GRN*-bvFTD.

As we expected, consistent with the widely acknowledged phenomenon that *C9orf72*-bvFTD or *GRN*-bvFTD typically exhibit TDP-43 pathology^34^, the gene sets associated with cortical thickness signature of both *C9orf72*-bvFTD and *GRN*-bvFTD showed significant enrichment for TDP-43 pathology-related DEGs. Conversely, *MAPT* variants predominantly lead to tau pathology, resulting in absence of TDP-43 pathology enrichment within the gene sets related to *MAPT*-bvFTD. We further examined the enrichment of tau pathology in the context of PSP. The gene sets associated with *C9orf72*- bvFTD and *GRN*-bvFTD did not yield significant results in this regard. This underscores the pathology-specific nature of these genes, aligning with the pathological presentations of these two genetic forms. However, genes associated with *MAPT*-bvFTD did not display significant enrichment for tau pathology. This could potentially be attributed to the fact that these tau-related genes were identified in PSP that link to 4R tauopathy, while FTLD-tau owing to different *MAPT* variants can result in 3R, 4R or mixed 3R/4R tauopathies^3^. Moreover, genes associated with *MAPT*-bvFTD were enriched in pathways not directly linked to nervous system, including “cholesterol biosynthesis and metabolism” and “enzyme activities and metabolism”. The identification of these overlapping genes between *C9orf72*-bvFTD or *GRN*-bvFTD and TDP-43 pathology, provides valuable insights into the shared molecular pathways that might underlie their potential involvement in disease pathogenesis.

We further delved deeper into the shared genes identified across *C9orf72*-bvFTD, *GRN*-bvFTD, and *MAPT*-bvFTD, indicating common molecular foundations among different genetic forms. Both *C9orf72*-bvFTD and *GRN*-bvFTD that linked to TDP-43 pathology^35^, their transcriptomic signatures were enriched in previously identified genes linked to TDP-43 positive neurons. These two genetic forms shared genes displaying consistent directionality, with those exhibiting either positive or negative correlations with cortical thickness in *C9orf72*-bvFTD showing the same direction (positive or negative) in *GRN*-bvFTD. *MAPT*-bvFTD that associated with tauopathy^35^, displayed more pronounced transcriptomic differences, since *MAPT*-bvFTD had shared genes with the other two forms, but with opposing directionality. These shared genes that are commonly expressed in regions more spared by cortical atrophy in *C9orf72*-bvFTD and *GRN*-bvFTD, and conversely, these genes are commonly expressed in regions more vulnerable to brain atrophy in *MAPT*-bvFTD. Thus, the shared transcriptomic signatures exhibited similar relationships with cortical remodeling due to pathogenic variants in *C9orf72* and *GRN*, while showed opposite relationships with cortical remodeling attributed to pathogenic variants in *MAPT*. Moreover, our focus further centered on several key genes within this shared gene pool. One prominent gene is *GNG2*, which was shared by *C9orf72*-bvFTD and *GRN*- bvFTD, in collaboration with other genes such as guanine-nucleotide binding proteins (*GNB2*, *GNB3*, *GNB4*, and *GNB5*), is known to be intricately involved in driving synapse-related pathways^36^. Another key gene is *SMARCD3*, in conjunction with *SMARCB1* and *SMARCC2*, encodes proteins belonging to the SWI/SNF family, which play crucial roles in regulating gene transcription through chromatin remodeling, thereby impacting neural development, proliferation and degeneration^37,38^. The *EVT6* gene presented in *GRN*-bvFTD and *MAPT*-bvFTD, might not exert a direct influence, but its interaction with RAS genes like *NRAS*, *KRAS*, and *HRAS* has implications for synaptic signaling and axon guidance. Additionally, the *CHRNA2* gene, along with *CHRNB2* gene, is associated with synaptic signaling processes. These findings illuminate key genetic factors contributing to the shared molecular mechanisms across genetic forms of bvFTD, enhancing our understanding of complex interplay within neurodegeneration.

There are several limitations to consider in future work. One limitation is the utilization of regional transcriptomic data from only six donors. We excluded data from the right hemisphere due to limited availability, potentially introducing biases related to asymmetric cortical atrophy and gene expression patterns across two hemispheres. This constraint reflects a broader challenge in the field, as acquiring a comprehensive whole-brain atlas of gene expression for all genes across all regions remains challenging. Secondly, we correlated this constitutive gene expression data with neuroimaging features derived from a different cohort. While it was assumed that regional gene expression serves as a conserved canonical signature and its generalizability extends far beyond these 6 brains^39,40^, this remains a noteworthy limitation that lies in the potential incomplete representation of our cohort by this regional gene expression atlas. In addition, the PLS regression model requires the same number of samples for predictor (e.g. transcriptome) and response (e.g. neuroimaging) variables, and in our case, the samples correspond to brain regions. Gene expression for each region was averaged across donors, while cortical thickness signatures were statistical differences between genetic and apparently sporadic bvFTD. Thus, sample size can influence the averaged gene expression and estimates of cortical thickness signatures, thereby influencing the correlations predicted by PLS regression model^41^. Given the relatively small sample size of *GRN*-bvFTD and *MAPT*-bvFTD groups, larger cohorts are essential for further validation. Additionally, the spatial colocalization between gene expression and cortical atrophy does not necessarily imply a causal relationship between them. The identified genes require validation through forthcoming animal experiments to assess their functions and impact.

In conclusion, we identified both disparate and shared transcriptomic signatures associated with cortical atrophy in bvFTD with pathogenic variants in *C9orf72*, *GRN*, and *MAPT*. The spatial distribution of gene expression may contribute to selective vulnerability of macroscopic neuroanatomy, giving rise to the heterogeneity observed in bvFTD. Our study provided evidence mapping genetic-related variance in cortical thickness to synaptic genes, circadian-related genes and dysregulated genes in TDP-43 pathology, linking molecular processes, cellular mechanisms, pathological manifestations, and macroscopic brain structure. These findings highlighted the disparate and shared molecular underpinnings of various genetic subgroups and their relationship with brain structural remodeling. With consideration of specific genetic variants involved, this approach can unravel the intricate molecular mechanisms driving the heterogeneous nature of genetic bvFTD. This knowledge might pave the way for precisely tailored strategies in diagnosis and potential treatments.

## Methods

### Study Oversight

The Institutional Review Board of the University of Pennsylvania gave ethical approval for this study and provided ethical oversight of the study conduct. All participants or their caregivers gave written informed consent prior to engaging in study procedures, in accordance with the Declaration of Helsinki.

### Participants

Participants were retrospectively selected from the Integrated Neurodegenerative Disease (INDD) database at University of Pennsylvania^42^, consisting of 173 individuals with bvFTD, and 172 age, sex matched healthy controls who self-reported a negative neurological and non-significant psychiatric history with a normal Mini-Mental Status Examination (MMSE) > 27. Individuals with bvFTD were clinically diagnosed based on published criteria^1^ and using a consensus procedure with > 2 clinical experts. Individuals with a concurrent diagnosis of amyotrophic lateral sclerosis (ALS), corticobasal degeneration, and progressive supranuclear palsy (PSP) were excluded.

Clinical and neuropsychological assessments were conducted at Penn Frontotemporal Degeneration Center. Neuropsychological test scores were obtained from the visit closest to MRI scan. Cognitive and behavioral changes were evaluated using MMSE and Philadelphia Brief Assessment of Cognition (PBAC)^43^.

### Genetic Screening

All participants underwent genetic evaluation including *C9orf72* repeat expansion testing, and sequencing and deletion/duplication analysis of *GRN* and *MAPT*. The genomic DNA was extracted from peripheral blood or frozen brain tissue samples^2^. The presence of a *C9orf72* repeat expansion was evaluated using a modified repeat-primed polymerase-chain reaction^44^. *GRN* and *MAPT* variants were identified from whole genome sequencing, whole exome sequencing, or targeted neurodegeneration sequencing panel datasets^42^. The bvFTD cases that were negative for *C9orf72* repeat expansions, or pathogenic variants in genes associated with ALS/FTD, were defined as sporadic bvFTD.

### Neuroimaging data acquisition and preprocessing

Structural T1-weighted MRI scans were acquired on a Siemens 3.0 Tesla scanner outfitted as a TIM Trio (*n* = 289) and subsequently as a Prisma Fit (*n* = 56). MRI scans were collected with magnetization-prepared rapid gradient-echo (MPRAGE) sequences as follows: 1) 3.0 Tesla Siemens TIM Trio scanner, 8 channel head coil, axial plane with repetition time (TR) ranging from 1620 ms to 1900 ms, echo time (TE) ranging from 3.09 ms to 4.38 ms, slice thickness = 1.0 mm or 1.5 mm, in-plane resolution = 0.98 × 0.98 mm. 2) 3.0 Tesla Siemens TIM Trio scanner, 64 channel head coil, sagittal plane with TR = 2200 ms or 2300 ms, TE ranging from 2.95 ms to 4.63 ms, slice thickness = 1.0 mm or 1.2 mm, in plane resolution = 1.0 × 1.0 mm. 3) 3.0 Tesla Siemens Prisma scanner, 64 channel head coil, sagittal plane with TR = 2400 ms, TE = 1.96 ms, slice thickness = 0.8 mm, in-plane resolution = 0.8 × 0.8 mm^45^.

Images were processed using the Advanced Normalization Tools software through standard preprocessing steps^46^, including N4 bias field correction, diffeomorphic and symmetric registration to ADNI-3 template, brain extraction, and segmentation into six-tissue classes using template-based priors. The custom template was in turn aligned to the MNI152 2009c Asymmetric T1-weighted template. The Schaefer 7-network atlas with 200 cortical parcels^47^ was warped from MNI152 space through the custom template to individual space. Then cortical thickness was extracted from each label. To identify gene-specific atrophy patterns, we employed a w-score procedure to normalize cortical thickness values by age, and sex for each individual with bvFTD, relative to the mean distribution of 172 healthy controls. We then calculated the difference in w-scored cortical thickness for each cortical region between genetic and apparently sporadic bvFTD using a linear model, while adjusting for disease duration. Since only the left hemisphere was used, a 100 × 1 vector of case-control t-statistics for regional cortical thickness was obtained.

### Transcriptomic data and preprocessing

We extracted transcriptomic data from six postmortem healthy adult brains in the publicly available AHBA database using abagen toolbox^48^. Only left hemisphere data were used, as right hemisphere samples were only available for two of the six donors. Transcriptomic data was processed according to previously described protocol including the following steps: probe-to-gene annotation, intensity-based filtering by a threshold of 0.5, probe selection by differential stability, donor aggregation in probe selection, matching tissue samples to the Schaefer 7-network atlas with 200 parcellation atlas^47^, and data normalization. Finally, we obtained expression levels for 15,633 genes at 100 cortical regions, resulting in a 100 × 15,633 regional transcription matrix.

### Transcription-neuroimaging association analyses

A partial least squares (PLS) regression model that implemented in R was used to investigate associations between cross-condition cortical thickness signatures in genetic bvFTD and brain regional gene expression. The regional transcription matrix (100 regions × 15,633 genes) was taken as predictor variables, and the regional cortical thickness signatures vector (100 regions × 1) was treated as response variables. The first PLS component (PLS1) consisted of the linear combination of weighted gene expression values that was most strongly related to the anatomical map of regional cortical thickness signatures, which was considered as the most important component. The variability and statistical significance of PLS1 weight for each gene was estimated by bootstrapping the response variables 5,000 times (resampling with replacement of the 100 cortical regions)^49^. The ratio of the weight of each gene to its bootstrap standard errors was used to calculate z scores and then transformed to *p-values* to evaluate the contribution of each gene to PLS1. Only genes with *p-values* < 0.05 were regarded as significant contributors to PLS1^50^. Significant genes with large positive or negative PLS1 weights were termed as PLS1+ or PLS1- gene sets.

### Functional enrichment analyses

To test whether these genes map to relevant biological pathways for biological interpretations, functional enrichment analyses were conducted on PLS1+ and PLS1- gene sets separately using Metascape^51^. Metascape identifies ontology terms that contain a statistically greater number of genes in common with the input gene list than expected by chance^51^. We used the 15,633 genes extracted from AHBA dataset as background list. Enriched terms were rendered by databases including Gene Ontology (GO), Kyoto Encyclopedia of Genes and Genomes (KEGG), Reactome, and Comprehensive Resource of Mammalian protein complexes (CORUM). Enriched terms with Benjamini-Hochberg false discovery rate (BH-FDR) corrected *q-value* < 0.05 were reported as significant terms. Metascape enrichment networks were further generated to show the intra-cluster and inter-cluster similarities of enriched terms using Cytoscape software. Moreover, the Metascape membership function was also used to flag genes that fell into terms related to different neurotransmitter systems (acetylcholine, dopamine, GABA, glutamate, and serotonin)^51^.

### Mapping to neurotransmitter maps

To further investigate the involvement of synaptic signaling in shaping brain structure, we related the different neurotransmitter receptor/transporter density maps to cortical thickness signatures of each genetic form of bvFTD. Maps of cortical distribution of neurotransmitter receptors/transporters were obtained from an assembled collection of PET scans of about 1,200 normal subjects (https://github.com/netneurolab/hansen_receptors)^52^. We selected 15 different receptors/transporters across five neurotransmitter systems, including acetylcholine, dopamine, GABA, glutamate, and serotonin. PET images were acquired using different imaging protocols for various radioligands, resulting in different measured values for each tracer image, such as binding potential and tracer distribution volume. For simplicity, we refer to all these measured values as density^52^. All images were also registered to the MNI152 2009c Asymmetric T1-weighted template, then parcellated to 200 regions according to the Schaefer 7-network atlas^47^. Group-averaged neurotransmitter density maps were then constructed.

We performed correlation analyses between neurotransmitter receptor/transporter densities maps and cortical thickness t-statistic map of each genetic form. We further considered correlations between variables by conducting dominance analysis to evaluate the contribution of each predictor (15 neurotransmitter receptors/transporters) in predicting outcome variable (cortical thickness signature) using multiple linear regression model^53^. The importance of each predictor was quantified using measures of total dominance and relative importance. Total dominance summarizes the additional contributions of a predictor to all possible subset models, and relative importance is expressed as a percentage of the total variance in the outcome variable that each predictor explains.

### Pathology-related gene enrichment analyses

We hypothesized that the significant PLS1 genes identified in individuals with *C9orf72-*bvFTD and *GRN-*bvFTD were associated with TDP-43 pathology, and genes associated with *MAPT*-bvFTD were more involved in tau pathology. To explore whether the significant PLS1 genes were enriched for genes related to TDP-43 pathology or tau pathology, we extracted the genes from previously published studies^12,13^. The genes related to TDP-43 pathology was defined by Liu and colleagues^12^. They fractionated and sorted for diseased neuronal nuclei from postmortem ALS-FTD human brains and identified 5,576 significantly differentially expressed genes (DEGs) that were associated with TDP-43 pathology. In terms of tau pathology, Allen and colleagues conducted correlation analyses between gene expression and a neuropathological latent trait of tau pathology in a PSP cohort to identify significant transcriptional associations. This resulted in identification of a list of 2,549 genes considered as tau pathology-related genes^13^.

For each genetic form, we drew random sets of genes the same size as the significant PLS1 gene set, and calculated the number of overlapping genes between the random gene set and the TDP-43 or tau pathology-related gene lists. These procedures were repeated 1,000 times to obtain a null distribution of overlapping gene numbers. The null hypothesis was that the observed number of overlapping genes between PLS1-/+ gene sets and pathology-related gene lists was not significantly higher than the overlap between random gene set and pathology-related gene lists. The *p-value* of enrichment for TDP-43 or tau pathology-related genes were estimated by comparing the observed overlapped gene number to the null gene counts distribution. Therefore, significant gene overlap indicated enrichment for genes involved in TDP-43 or tau pathology.

### Shared transcriptional signatures across genetic forms

To explore the shared transcriptional signatures across different genetic forms, we focused on determining the extent of gene overlap between each pair of genetic forms by calculating the number of overlapping PLS1 genes present in each combination. These overlapping genes were then analyzed by conducting protein-protein-interaction (PPI) network analysis to predict functional partner genes provided by STRING database, encompassing both physical and functional associations^54^. Moreover, we proceeded to investigate the expression patterns of these overlapping genes across human tissues and cell types by leveraging data from the Human Protein Atlas (HPA) and genotype-tissue expression (GTEx) databases^55^. To portray gene expression levels, we extracted normalized transcript per million (nTPM) values, providing a quantitative representation of gene’s activity.

## Data Availability

All data used in this study are available upon reasonable request and approval from the Penn Neurodegenerative Data Sharing Committee. Requests may be submitted using a webform request: https://www.pennbindlab.com/data-sharing.

## Acknowledgements

This work was supported by NIH funding (P30 AG072979, P01AG066597, R01NS109260), Penn Institute on Aging, Robinson Family Fund, Peisach Family Fund for FTD Research, DeCrane Fund for Primary Progressive Aphasia, and Arking Family Fund. Jacob W Vogel acknowledges funding from the NIH (T32MH019112) and the SciLifeLab & Wallenberg Data Driven Life Science Program (grant: KAW 2020.0239). Jeffrey S. Phillips was supported by NIH grant (R01-AG054519, K01-AG061277).

## Author contributions

T.S., J.W.V. and C.T.M. conceived and designed the study. V.V.D., L.M. and D.J.I contributed to acquisition and analysis of clinical data. L.D. contributed to genetic data collection and testing. J.S.P. contributed to acquisition and analyses of MRI data. E.R.S. and E.B.L. helped with transcriptional data analyses. T.S. and C.T.M. wrote and reviewed the paper. All authors edited the paper.

