## Supplementary material for "Disparate and shared transcriptomic signatures associated with cortical atrophy in genetic bvFTD": Figure S1-S4

- **Figures S1-S4**

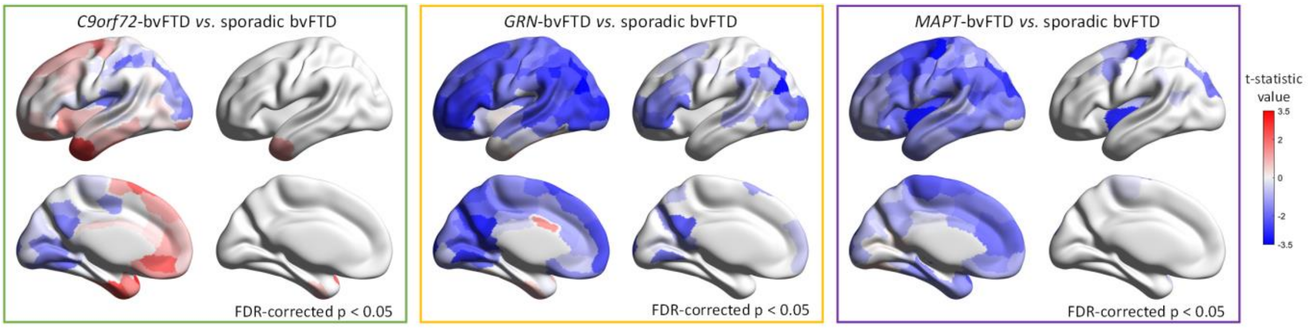


**Figure S1. Comparison of cortical thickness between genetic and apparently sporadic bvFTD.**

For each panel, the brain heatmap on left showing the t-statistic values of all brain regions, and the brain heatmap on right showing the regions with significant differences compared to apparently sporadic bvFTD.

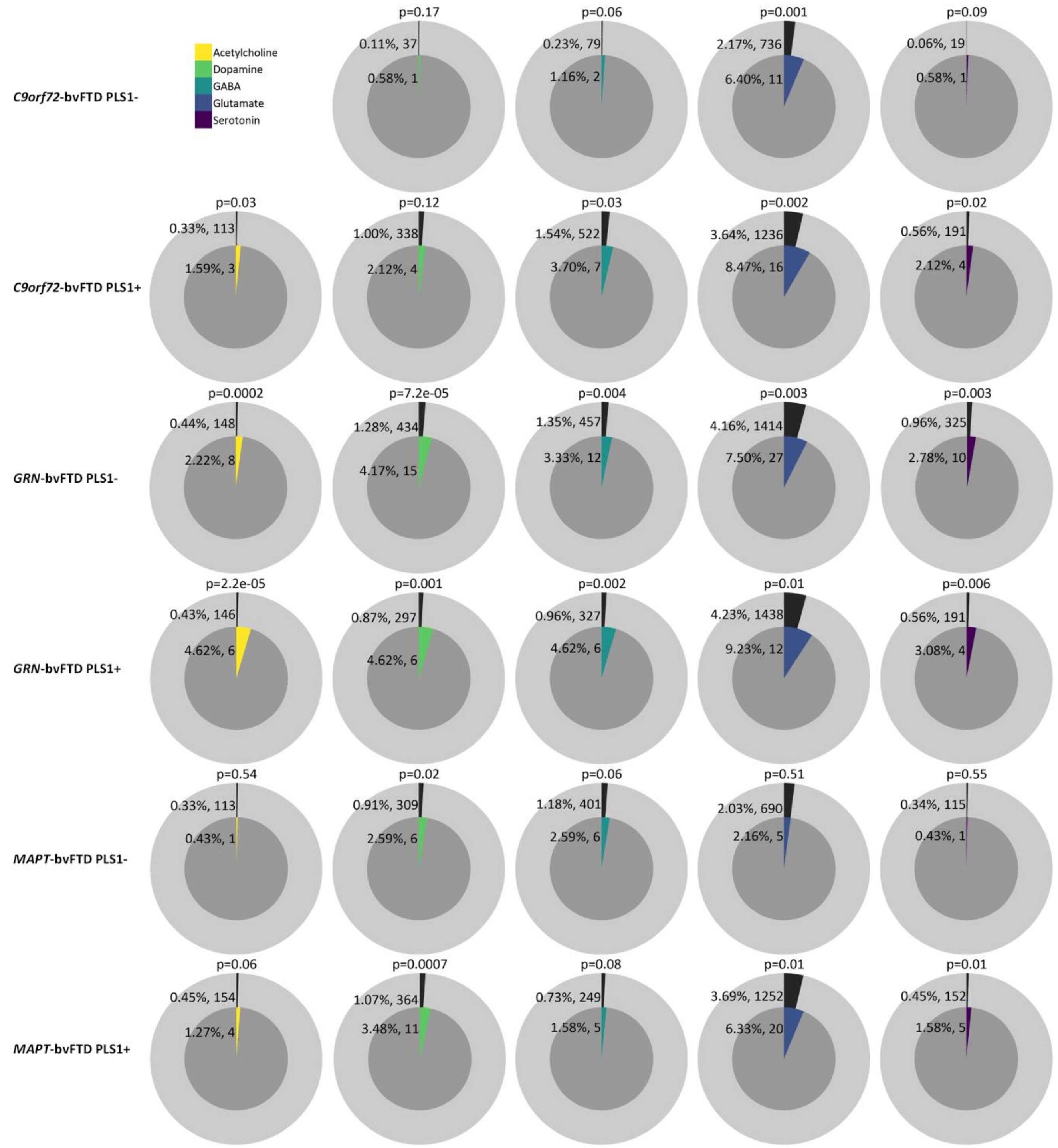


**Figure S2. Metascape membership analysis for PLS1-/+ gene lists associated with cortical thickness signatures of each genetic form of bvFTD.**

Membership search was conducted for terms related to a cholinergic, dopaminergic, GABAergic, glutamatergic, and serotonergic neurotransmitters. The outer ring of each pie represents the number and percentage of genes that are members of selected ontology terms, the inner ring shows the number and percentage of genes in the PLS1-/+ gene lists that are members of selected ontology term. The *p values* at the top of each pie indicates whether the selected term is statistically enriched in the input gene list.

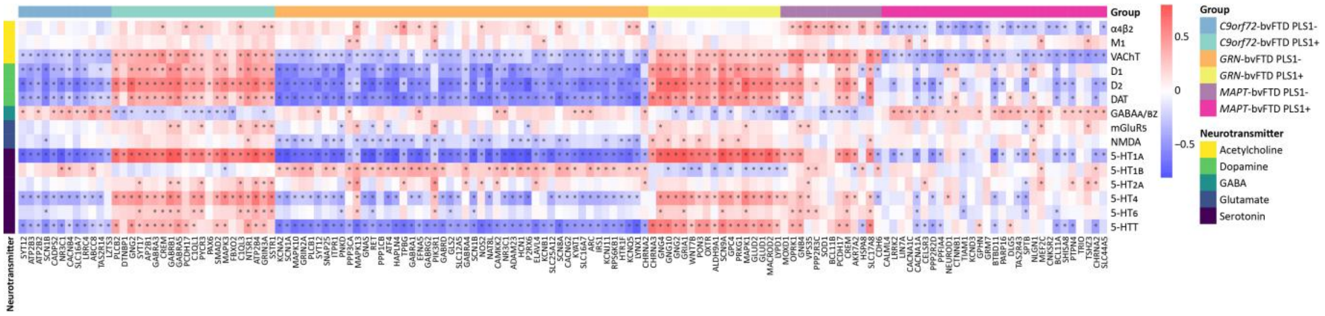

**Figure S3. Correlations between the gene expression maps and neurotransmitter receptor/transporter density maps.**

* indicates that the correlation is significant.

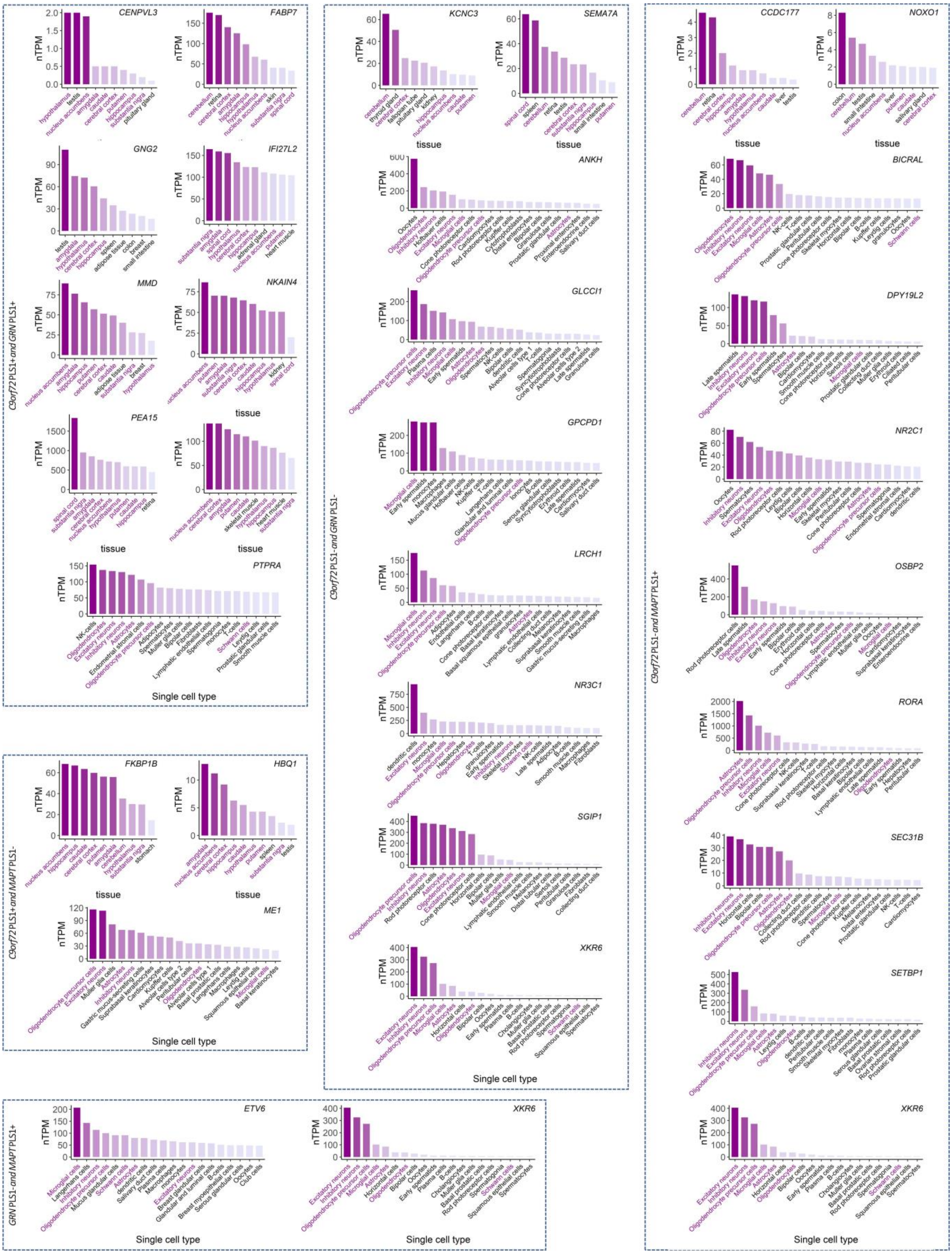


**Figure S4. Expression patterns of genes in multiple tissues and cell types.**

Only genes specifically enriched in either brain tissues or neural cells were shown.
